# Verbosity with retelling: narrative discourse production in temporal lobe epilepsy

**DOI:** 10.1101/2022.09.01.22279515

**Authors:** Fiore D’Aprano, Charles B. Malpas, Stefanie E. Roberts, Michael M. Saling

**Affiliations:** Melbourne School of Psychological Sciences, The University of Melbourne, Australia; Department of Neurology, The Royal Melbourne Hospital, Australia; Department of Neurology, Alfred Health, Australia; CORe, Department of Medicine, The University of Melbourne, Australia; Department of Clinical Neuropsychology, The Austin Hospital, Australia

**Keywords:** discourse, circumstantiality, verbosity, language, temporal lobe epilepsy, pragmatics

## Abstract

The conversational language of individuals with temporal lobe epilepsy (TLE) is circumstantial. The micro- and macrolinguistic underpinnings of this disturbance in narrative discourse, and the role of epilepsy and cognitive variables warrants exploration. We examined the elicited narrative output of 15 surgically-naïve individuals with TLE and 14 healthy controls. To replicate and extend Field and colleagues’ (2000) work, participants were shown an eight-frame cartoon Cowboy Story from Joanette and colleagues (1986) and were asked to produce five immediately consecutive elicitations of the narrative. Following transcription and coding, detailed multi-level discourse analysis demonstrated a typical pattern of compression across repetitions in controls. They produce increasingly concise and coherent output, reflective of a refined mental representation of the narrative, while individuals with TLE fail to do so. The narratives produced by individuals with TLE do not compromise the essential story components, although they are less informative overall: producing fewer novel units, and introducing more content that is repetitive, extraneous, and does not progress the narrative. Their narratives are ultimately less fluent, less cohesive, and less coherent relative to controls. Change across trials suggests that there are significant group by trial interactions in sample length, spontaneous duration, and total statements, which are not explained by seizure burden, age, or lexical retrieval deficits among those with TLE. These findings replicate the pattern of findings previously identified by Field and colleagues (2000), with novel insights into the macrolinguistic disturbances that characterise their narrative discourse over sequential repetitions. These findings suggest that individuals with TLE do not benefit from repeated engagement with a narrative in the same way that neurologically normal individuals do. We conclude that disturbances to social cognition and ultimately pragmatics in TLE might underpin inefficiencies in their communication.

## Introduction

Individuals with temporal lobe epilepsy (TLE) have well-documented language disturbances at the single-word level. Conversationally, their pedantic, repetitive, and highly detailed output style is termed ‘circumstantiality’ (Bear, Levin, Blumer, Chetham, & Ryder, 1982; Benson, 1991; Geschwind, 1977). This pattern of verbosity is observed clinically, and since the original formulation of Bear and Fedio (1977), is now considered a phenomenon attributable to subtle interictal disruptions of neurolinguistic functions rather than a personality feature (Field et al., 2000; Rao et al., 1992). Circumstantiality in TLE might serve to compensate for lexical retrieval deficits which impact fluency (Brandt, Seidman, & Kohl, 1985; Busch et al., 2013; Mayeux, Brandt, Rosen, & Benson, 1980), suggesting that lexical-syntactic dysfunction directly relates to macrolinguistic impairments. Different linguistic contexts and challenges produce different psycholinguistic phenotypes in TLE, where output from a single spontaneous elicitation is characterised by verbosity to a greater extent than output in a structured context when compared to controls (D’ Aprano, Malpas, Roberts, & Saling, 2022 pre-print). Where structure is imposed on discourse there is a semantically limited and largely invariable scope of lexical and propositional choices (Smith, Heuerman, Wilson, & Proctor, 2003). When produced by neurologically normal individuals, the structured elicitation of impersonal narratives are not expected to deviate substantially from the core content. On a single elicitation of a six-frame narrative, individuals with TLE were not verbose compared with controls (Bell et al., 2003). Instead, they produced a similar number of words and spoke for a similar length of time, but were less fluent overall, producing more noncommunicative fillers, frequent abandonment of trains of thought, and higher incidence of repetitions relative to controls (Bell et al., 2003).

When a narrative is repeated, content becomes increasingly familiar and processing demands associated with initial planning and formulation decrease (Field et al., 2000; Goldman-Eisler, 1968). Re-telling aids the refinement of a discourse representation (Bloom, 1994) by minimising redundant or extraneous content to produce an increasingly succinct and relevant story. This is seen among controls where narratives repeatedly elicited from cartoon images become progressively concise. Their word count and duration of output reduces while their fluency increases (Field et al., 2000; Goldman-Eisler, 1968). These are markers of efficient discourse processing, and the refinement over repetitions is termed ‘the compression effect’ (Field et al., 2000). Unlike controls, individuals with left TLE fail to compress their discourse over repetitions, and instead become more verbose, that is, they speak for longer and use more words to convey increasingly familiar content (Field et al., 2000). Despite this,individuals with TLE are no less fluent than controls and tend to increase their fluency across repetitions (Field et al., 2000; Howell, Saling, Bradley, & Berkovic, 1994). Their verbosity appears unrelated to psychometrically defined impairments at the single-word level (Field et al., 2000). The failure to compress discourse therefore reflects inefficiencies in producing and refining a mental representation of a narrative, rather than lexical processing deficits (Field et al., 2000; Saling et al., 2014). These findings suggest that micro- and macrolinguistic components are dissociable in TLE, being those that relate to lexical-syntactic and suprasentential processes, respectively.

Metrics of verbosity often examine output volume, rather than the composition of output. From early descriptions of circumstantiality in TLE, repetitiveness and high levels of detail also contribute to the overall syndrome. These macrolinguistic disturbances, including tangential or irrelevant details, and stereotyped phrases, reduce the informativeness of output and impact global coherence (Marini, 2012). Coherence relates to suprasentential organisation, that is, how content is connected thematically to reach a goal (Agar & Hobbs, 1982; Glosser & Deser, 1991). Poor coherence in the face of decreasing processing demands reflects an inefficiency in refining a mental representation and impacts discourse quality. These impairments in informativeness and coherence are akin to those identified in healthy aging (Glosser & Deser, 1991; Juncos-Rabadán, Pereiro, & Rodríguez, 2005; Marini, Boewe, Caltagirone, & Carlomagno, 2005). While younger participants benefit from repetition and progressively produce concise and cohesive narrative discourse, the same is not seen among the elderly (Saling, Laroo, & Saling, 2012).

Using functional magnetic resonance imaging to examine discourse compression in healthy controls, Lillywhite and colleagues (2010) highlight the role of the middle frontal gyrus bilaterally and right inferior parietal region in re-listening to narratives. They suggest that repeated engagement with the narrative progressively recruits a broader network, beyond regions fundamentally involved in comprehension and production. Their findings also implicate the right inferior parietal region in representing discourse, with local blood-oxygen- level-dependent (BOLD) signal correlated with the ‘word count’ metric of discourse compression in the Cowboy Story task. These findings form part of an emerging literature regarding the role of the right hemisphere in discourse processing (Abusamra, Côté, Joanette, & Ferreres, 2009; Johns, Tooley, & Traxler, 2008; Xu, Kemeny, Park, Frattali, & Braun, 2005). Patients with right hemisphere damage, particularly with anterior lesions, appear to have intact microlinguistic function. Impairments emerge at the macrolinguistic level when considering global coherence and overall informativeness, producing more tangential content (Abusamra et al., 2009; Marini, 2012). Right frontal regions, as part of complex networks, appear to play a role in organising information coherently in a narrative discourse (Marini, 2012). Individuals with right TLE have similar discourse processing deficits in narrative and conversational contexts in the absence of lexical or syntactic deficits (Lomlomdjian et al., 2017), which might be subserved by disrupted projections from temporal to frontal regions in TLE (Devinsky, 2005; Lieb, Dasheiff, Engel, Genton, & Genton, 1991; Stretton & Thompson, 2012). In light of broader network dysfunction in TLE, disruptions to discourse, as a higher cortical function, are unlikely lateralised as these processes rely on a multitude of disparate aspects of cognition including planning and working memory (Coelho, Grela, Corso, Gamble, & Feinn, 2005; Shulman, 2000).

The present study aimed to replicate and extend the seminal work by Field and colleagues (2000) by expanding the number of elicitations of a narrative and examining discourse from a multi-level perspective. We aimed to discern what contributes to language disruptions at micro- and macrolinguistic levels to further our understanding of circumstantiality in narrative discourse among patients with TLE. We hypothesised that healthy controls would produce increasingly concise output, manifesting as a reduction in total output, greater fluency, and improved cohesion and coherence over successive repetitions. We anticipated that among individuals with TLE there would be impairments of fluency, cohesion, and coherence and a failure to compress discourse over repetitions. These disturbances were hypothesised to reflect inefficiencies in their capacity to refine a mental representation of the discourse.

## Methods

### Participants

This study included 29 participants, 15 focal unilateral TLE comprising 10 left TLE (5 mesial, 3 neocortical, 2 non-lesional) and 5 right TLE (4 mesial, 1 neocortical), and 14 healthy controls. Inclusion criteria were: English as a first language, a diagnosis of drug-resistant TLE when recruited (Kwan et al., 2010), no prior neurosurgical resection, full scale IQ >70, no reported history of substance-related and addictive disorders, no formally diagnosed psychiatric disorders, and no current major psychiatric episode (e.g., psychosis). None of these individuals were receiving additional treatments for the control of seizures (e.g., vagal nerve stimulation) and none had a history of developmental language disorder or other neurological condition (e.g., stroke). To localise seizure focus, these participants underwent multi-day video electroencephalography (Video-EEG) in either The Royal Melbourne Hospital or the Alfred Hospital in Melbourne, Australia. Diagnostic decisions are made by a comprehensive team comprising neurologists, epileptologists, neurophysiologists, psychiatrists, and neuropsychologists in accordance with criteria defined by the International League Against Epilepsy (ILAE; (Engel Jr, 2006)). Consensus based on seizure semiology, video-EEG, magnetic resonance imaging (MRI), positron emission tomography (PET) and inter-ictal single-photon emission computer tomography (SPECT) provided unambiguous diagnoses with a localised seizure focus. By assessment, three TLE participants reported no seizures in the preceding 12 months on their current anti-seizure medication (ASM) regimen (Kwan et al., 2010). Analyses were performed both with and without these drug-responsive participants to determine whether their inclusion in the sample was a robust choice. We found no difference in key outcome measures and these individuals were subsequently retained in the sample. Their reduced epilepsy burden at that point in time is reflected in the 13-point Seizure Frequency Rating (So et al., 1997)—a composite metric considering seizure frequency, type, and ASM usage. Healthy controls comprised family members or partners of TLE participants, and where necessary were recruited from the community via convenience sampling to age-, education-, and sex-match to those with TLE. Demographic characteristics for these groups are provided in Table 1, who were broadly comparable on variables of interest.

**Table 1.** Sample characteristics

Sample characteristics
| Variable | Control<br>(n = 14) |  | TLE<br>(n = 15) |  | p | Effect size |
| --- | --- | --- | --- | --- | --- | --- |
|  | Median (Q1, Q3) | Range | Median (Q1, Q3) | Range |  |  |
| Age [years] | 51.50 (26.00, 55.50) | 50 | 43.00 (32.00, 53.50) | 44 | 0.96 | 0.01 |
| Education [years] | 15.50 (12.00, 18.00) | 9 | 15.00 (12.00, 17.00) | 15 | 0.58 | 0.12 |
| Estimated IQ<sup>a</sup> | 107.75 (97.63, 111.88) | 41 | 102.50 (95.25, 107.00) | 50.50 | 0.20 | 0.29 |
| Age at Diagnosis [years] | N/A | N/A | 25.00 (20.00, 43.50) | 50 |  |  |
| Epilepsy duration [years] | N/A | N/A | 7.00 (3.00, 17.50) | 41.50 |  |  |
| Seizure Burden Rating | N/A | N/A | 6.00 (2.00, 7.00) | 8 |  |  |
| Last seizure [weeks] | N/A | N/A | 8.00 (4.00, 40.00) | 75 |  |  |
| Current ASMs | N/A | N/A | 2.00 (1.00, 2.00) | 3 |  |  |
| Laterality, Left [n, %] | N/A | N/A | 10 (67) |  |  |  |
| Mesial focus [n, %]<sup>^</sup> | N/A | N/A | 9 (60) |  |  |  |
| Sex, Female [n, %] | 7 (50) |  | 7 (47) |  | 0.86 | 0.03 |
| Handedness, Right [n, %] | 12 (86) |  | 12 (80) |  | 0.68 | 0.08 |

*Note.* TLE = Temporal Lobe Epilepsy; ASM = Anti-seizure Medication. For continuous variables, group differences were computed using Mann-Whitney U test, effect size reported is the rank biserial correlation coefficient. <sup>a</sup>Suggests that data does not violate assumptions of normality on Shapiro-Wilk. For discrete variables, percentages of each group reported, group differences computed using X<sup>2</sup> test of independence, effect size reported is phi-coefficient. Estimated IQ reflects a composite between TOPF scaled score and WASI-II FSIQ-2 score. Seizure frequency calculated on 13-point rating scale (0-12) considering ASM usage, frequency, and type of seizure (So et al., 1997). <sup>^</sup>Of these individuals 5 had a diagnosis of left TLE. Non-mesial cases comprise 4 neocortical (3 left, 1 right), and 2 non-lesional (left seizure focus).

This multi-site study received ethical approval from the Melbourne Health Human Research Ethics Committee in accordance with ethical standards of the 1964 Declaration of Helsinki. All participants provided written informed consent.

### Neuropsychological Assessment and Discourse Elicitation

This study involved neuropsychological and language assessment (Appendix A), conducted by a single registered psychologist. Given the COVID-19 lockdown conditions in Melbourne, Australia at the time of collection, these assessments were completed via telehealth. Metrics of lexical retrieval included the Boston Naming Test (BNT) (Kaplan, Goodglass, & Weintraub, 1983), the Controlled Oral Word Association Test (COWAT) (Strauss, Sherman, & Spreen, 2006), the Auditory Naming Test (ANT) (Hamberger & Seidel, 2003) with minor modifications to suit the Australian lexicon, and the Verb Generation Task (VGT) developed to examine verb retrieval (Appendix A).

Participants were shown an eight-frame cartoon story called “Cowboy Story” (Joanette et al., 1986) via screen-share which remained in front of participants for the duration of their description to minimise any memory contribution. The story was elicited on five immediately consecutive occasions. On the first trial, participants were encouraged to take enough time to understand the cartoon and were prompted to “tell me in as much detail as you can everything you see happening in this story, tell it to me as though I cannot see the cartoon”. For all subsequent trials (total five), participants were asked to “tell me the same story again now please”. Participants were not interrupted for the duration of their output and were required to make a definitive statement to indicate that they had completed each trial; for example, “I’ m finished”. The researcher minimised verbal and non-verbal participation throughout the elicitations.

### Recording and transcription

Audio output was recorded from the Zoom session (Zoom Video Communications Inc, 2016) then manually transcribed verbatim and segmented by a single researcher within four weeks of the file being obtained. Consistent with methodology applied by Stein and Glenn (1979) and Trabasso and van den Broek (1985), statements were segmented so that a single statement refers to a predicate and its corresponding arguments. This provides a proposition- based extraction of content and analysis of coherence (Davis, O’ Neil-Pirozzi, & Coon, 1997), rather than by communicative unit (C-unit). Pause lengths in grammatical and non-grammatical junctures were manually extracted from Audacity^®^ software. Sample lengths refer to the total number of completed words within a sample, excluding words that fill pauses such as “um” “uh” (Nicholas & Brookshire, 1993; Stockbridge et al., 2021).

### Discourse variables and coding practices

Based on multiple models of discourse production and examination in clinical populations, a set of discourse variables were selected to conduct a multi-level discourse analysis (Sherratt, 2007). These relate to key aspects of linguistic analysis, at both the micro- and macrolinguistic level, being lexical-syntactic and suprasentential, respectively. A description of all nodes coding is presented in Appendix B. Using NVivo 12 software, transcripts were coded by a single researcher, with expertise in cognition and linguistics, who was blind to participant characteristics other than date of data acquisition. Each transcript was coded twice by the same reviewer, with an intra-rater agreement of 95% across both coding occasions. Where discrepancies were identified, the researcher re-considered the coding criteria and produced a final decision. Any ambiguities in the description of criteria for coding to nodes were clarified.

### Coding agreement

To establish inter-rater agreement, a second expert researcher blindly coded a randomly-selected subset of the total transcripts. In line with other similar analyses of discourse, this was determined to be 12.5% (Sherratt, 2007) and included a total of five transcripts. Inter-rater agreement was determined on a point-by-point basis in terms of the specific node to allocate as well as appropriate statement segmentation. The second researcher was blinded to participant characteristics and had access to the complete, disambiguated codebook for this process. For nodes, percentage of agreement was 89%, while for the segmentation of statements, there was 96% agreement—both surpassing the minimal accepted requirement level of 80% (Kazdin, 1982). Coding discrepancies were discussed between among the reviewers and resolved via consensus. Once again, any ambiguities in the description of nodes were resolved and their coding was updated (Appendix B).

### Statistical Analyses

For sample characteristics and neuropsychological measures, analyses were conducted using Jamovi software (Jamovi, 2021) to compute group-specific measures of central tendency, to assess group differences, and to calculate partial correlations. Many of the data were skewed. To be conservative, non-parametric tests (Mann-Whitney U test) have been applied across all analyses for the purpose of uniformity. Data that did not violate assumptions of normality are indicated in table notes. Contingency tables (X^2^ test of independence) were used for categorical variables. The rank biserial correlation (RBC) was used as a non-parametric estimate of effect size, reflected as small 0.1 < 0.3 < 0.5 large. Partial correlations were used to examine the relationships between core discourse variables at Trial 5, demographic, and seizure characteristics while controlling for Age. These are reported as Spearman’ s rank correlation coefficients.

To ensure that analyses were not sensitive to an artifact such as group, we ran the analyses with left and right TLE separately and then together. These analyses suggested no difference in discourse outcomes between left and right TLE or relative to controls when individuals with TLE were considered as separate groups or a single group. Consistent with the notion that high-level language functions are not lateralised, all individuals with TLE were subsequently treated as a single group. This methodological point is further addressed in the discussion. To account for Type I error, a false detection rate (FDR) of 0.05 was applied to primary analyses (Benjamini & Hochberg, 1995). General linear mixed models (GLMMs) were estimated using the GAMLj package for Jamovi software, with group as factor, participant as cluster variable, and trial as covariate.

## Results

### Sample characteristics

Individuals with TLE and controls were comparable across most demographic characteristics and many aspects of neuropsychological function (see Table 1 and Appendix C, Table 1). Individuals with TLE reported higher rates of depressive symptomatology than controls. The TLE group broadly demonstrated a lexical retrieval deficit, and word findings difficulties were reported to be more frequent and more distressing than controls. While this was not reflected in the total number of correct items on measures of lexical retrieval, word finding difficulties manifested as *delays* in word retrieval: increased mean response time latencies and increased tip-of-the-tongue (TOT) states, that is, responding >2000ms post-stimulus *or* requiring phonemic prompting. This is true for BNT and ANT metrics of total TOT states, TOT states as a proportion of all responses, and for mean response latencies. Compared to controls, individuals with TLE also demonstrated longer latencies on VGT and produced fewer words within a semantic category.

### The TLE phenotype for repeated narrative

Group differences on discourse metrics at each trial are presented in Appendix C, Table 2. On the initial telling of the narrative (Trial 1), individuals with TLE are significantly less fluent than controls and have more cohesion disruptions. After five sequential repetitions, there are significant differences between controls and individuals with TLE at micro- and macrolinguistic levels which can be visualised in Figure 1. At the microlinguistic level, those with TLE have a slower production rate, produce more fluency disruptions, including more non-grammatical pauses and hesitations overall, and a longer duration of pauses. At the macrolinguistic level, those with TLE have more cohesion disruptions, speak for a longer duration, and use more other referents (ambiguous, incomplete, or missing). With regards to informativeness, both groups are comparable in the number of core story propositions (Appendix C, Table 2) produced across all trials. Those with TLE produce fewer task-on novel units and more non-progression units than controls and produce proportionately more statements that do not progress the narrative content relative to novel statements, that is, they are less informative in the content they communicate.

**Figure 1.**
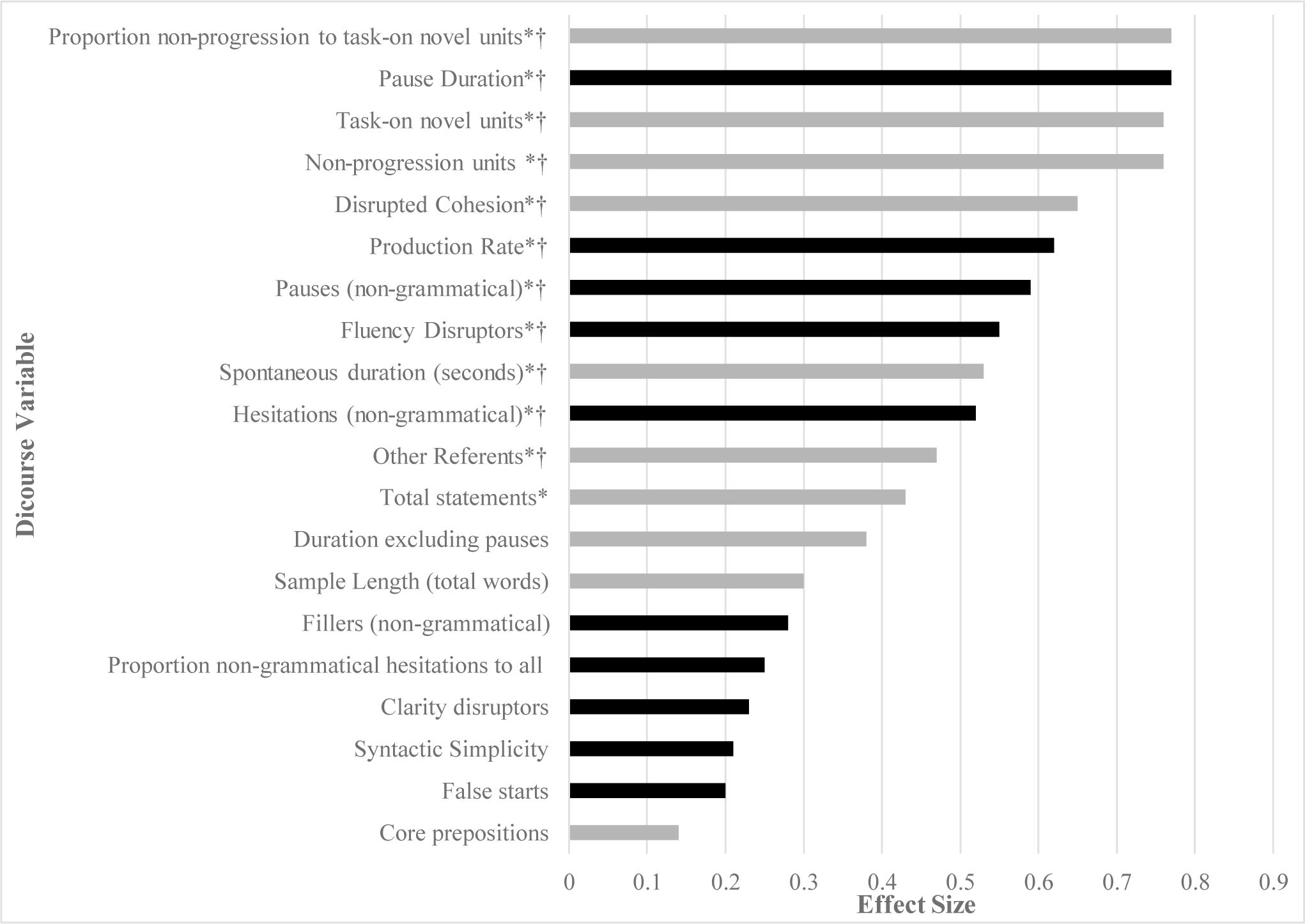
Mean differences on discourse variables relative to sample length between TLE and controls for Cowboy Story task at Trial 5, represented as absolute value effect size (rank biserial correlation, small 0.1 < 0.3 < 0.5 large), * = *p* < .05, † = significance holds on false detection rate (FDR) correction. Microlinguistic features are represented in black, macrolinguistic features are grey.

### Change over trials

Focusing on core verbosity and informativeness metrics, distinctions between individuals with TLE and controls emerge around the Trial 2 to 3 mark and we see a continuation and oftentimes deepening of this effect over additional repetitions. When adjusting for age, production rate at Trial 5 negatively correlates with disease duration (*r* = - 0.49, *p* = .008) and seizure burden (*r* = -0.53, *p* = .004), which in and of themselves are correlated (*r* = 0.59, *p* = .001); seizure burden also positively correlates with spontaneous duration (*r* = 0.57, *p* = .002); sample length (*r* = 0.44, *p* = .019); total statements (*r* = 0.43, *p* = .022); and ratio of non-progression to novel content (*r* = 0.59, *p* = .001). For these core variables, when adjusting for age, there are no other significant associations with demographic or seizure characteristics at Trial 5.

When modelling change over time, covarying for age and seizure burden did not impact the fixed effects for either sample length, spontaneous duration, total statements, production rate, or proportion of non-progression to task-on novel units. As such the models are reported without these covariates.

#### Sample length

Controls tend to tell the story in fewer words over repetitions, with a mean reduction of 48.71 words (*SD* = 51.89). While individuals with TLE are not frankly verbose, they do not reduce their output over retellings to the same extent as controls, with a mean decrease of 14.87 words (*SD* = 56.42) by Trial 5. A linear mixed model demonstrates a significant group by trial interaction *F*(1, 114) = 5.67, *p* = .019; a significant effect of trial, *F*(1, 114) = 10.94, *p* = .0013; and no significant effect of group, *F*(1, 27) = 1.25, *p* = .273. Figure 2a suggests that the significant interaction effect is predominantly driven by the decrease in sample length among controls.

**Figure 2.**
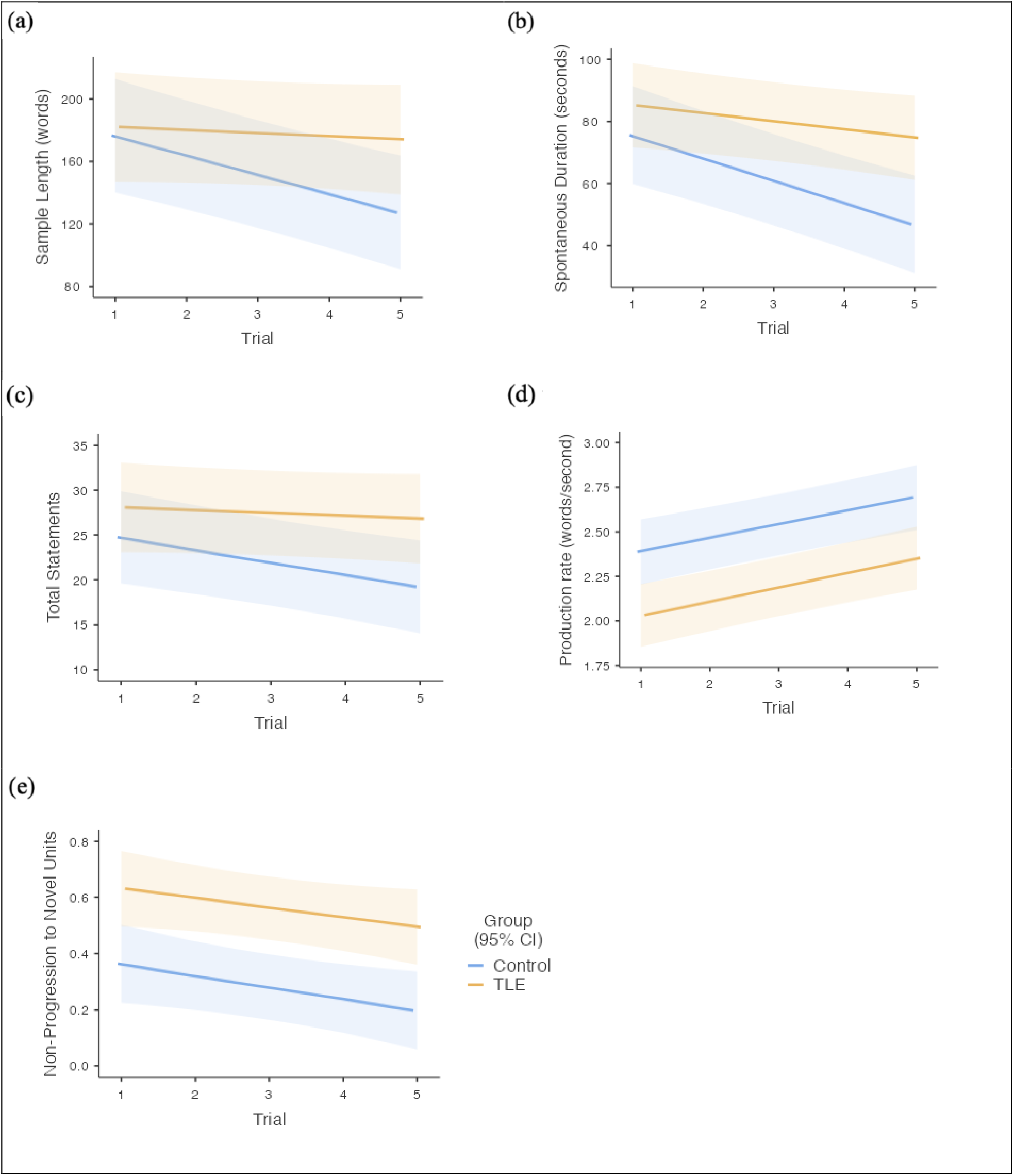
General linear mixed model effects plots for mean (a) sample length (words); (b) spontaneous duration (seconds); (c) total statements; (d) production rate (words/second); (e) proportion of non-progression to task-on novel units across trials in TLE and controls. Errors bars represent 95% confidence interval.

#### Spontaneous Duration

Over repetitions, output duration decreases for both groups. A linear mixed model suggests a significant group by trial interaction, *F*(1, 114) = 5.41, *p* = .022; a significant effect of trial *F*(1, 114) = 31.68, *p* < .001, but no significant effect of group, *F*(1, 27) = 4.19, *p* = 0.051. Figure 2b suggests that the significant interaction effect is predominantly driven by the decrease in duration among controls, where their magnitude of change across trials is significantly greater than TLE (*p* = 0.029, *RBC* = 0.45).

#### Total Statements

There is a significant group by trial interaction effect for the number of total statements produced, *F*(1, 114) = 5.11, *p* = .025; a significant effect of trial, *F*(1, 114) = 6.95, *p* = .0095; and no significant effect of group, *F*(1, 27) = 2.76, *p* = .108. Figure 2c suggests that the significant interaction effect is predominantly driven by the incremental reduction in total statements produced by controls, while those with TLE demonstrate more stability.

#### Production Rate

Both groups significantly increase their production rate across repetitions, see Figure 2d. A linear mixed models demonstrates a significant effect of trial, *F*(1, 114) = 48.33, *p* < .0001; a significant effect of group, *F*(1, 27) = 8.85, *p* = .0061; and no significant group by trial interaction effect, *F*(1, 114) = 0.04, *p* = .849. While the magnitude of improvement in fluency across trials is comparable between groups (*p* = .956, *RBC* = 0.02), those with TLE are less fluent to begin with and remain consistently less fluent than controls overall.

#### Proportion Non-progression to Novel Units

While both groups reduce the proportion of non-progression content across trials, those with TLE retain a consistently higher proportion of non-progression units to task-on novel units than controls, see Figure 2e. A linear mixed model demonstrates a significant effect of group, *F*(1, 27) = 12.71, *p* = .0014, a significant effect of trial, *F*(1, 114) = 7.45, *p* = .0073, but no significant group by trial interaction, *F*(1, 114) = 0.06, *p* = .800.

### Relationship to lexical retrieval

To examine whether these significant group by trial interaction effects for sample length, spontaneous duration and total statements might be due to a lexical retrieval deficit among individuals with TLE, the linear mixed models were repeated to separately consider BNT raw score, BNT TOT states, and COWAT performance as covariates. There were no changes in the pattern of significance with any of these variables.

## Discussion

Examining impersonal discourse via repeated narrative elicitation allows us to consider how dysfunctional discourse in TLE paradoxically emerges in the face of reduced processing demands. While microlinguistic impairments in this population are reasonably well-established, the macrolinguistic disturbances and their underpinnings remain unclear. This work extends that of Field and colleagues (2000) by examining additional elicitations using detailed multi-level discourse analysis procedures. This provides a unique opportunity to examine the profound and pervasive impact of TLE on discourse which penetrates multiple levels of neurolinguistic function. This study deals with the disturbances to fluency, cohesion, and coherence which characterise TLE.

Consistent with the findings of Field and colleagues (2000), relative to controls individuals with TLE failed to compress their discourse across consecutive narrative repetitions, although we did not see the incremental effect of verbosity in TLE that they described. The relationship is similar for sample length, spontaneous duration, and number of total statements, where there is relative stability across trials in TLE, and a decrement among controls who demonstrate a typical compression effect. Repetition is presumed to encourage refinement of the narrative by progressively overcoming the demands of newly planning and formulating discourse (Bloom, 1994; Goldman-Eisler, 1968). This process initially involves controlled processing to form an appropriate lexical space. Over repetitions, as this lexical space becomes more defined and delimited, content becomes more succinct and relevant via processes of deletion and generalisation (Goldman-Eisler, 1968). The absence of this benefit from repetition in TLE, in a setting of decreasing processing demands, reflects inefficiencies in discourse processing and refinement (Field et al., 2000). While discourse compression comprises high-level language processes, it is worth considering the influence of more fundamental aspects of discourse production which will be subsequently discussed.

Field and colleagues (2000) posited that lexical retrieval deficits among individuals with TLE do not account for macrolinguistic disturbances to output volume. This is supported by our current findings which suggest that metrics of lexical retrieval have no bearing on the interaction effects we observed for output volume. While lexical processing deficits do not purely account for discourse level impairments, microlinguistic processes still appear to influence discourse production through disruptions to fluency. Fluency disruptions are evident in TLE from the first trial, particularly in the form of pauses and hesitations at non-grammatical junctures. This is consistent with previous studies (e.g., Bell et al., 2003). Under these structured circumstances, the scope of discourse is largely invariable and the semanticolexical and propositional space is limited (Smith et al., 2003). Individuals with TLE are therefore expected to perform comparably to controls on macrolinguistic features, but demonstrate microlinguistic disturbances at a sentential level which impacts their fluency (Bell et al., 2003;D’ Aprano et al., 2022 pre-print). Fluency disruptions and lengthy pause durations reduce with retelling in healthy individuals (Goldman-Eisler, 1968), but as our data show, persist in individuals with TLE. To the extent that these disruptions reflect poor self-monitoring and planning of output (Goldman-Eisler, 1968), they are likely to impact discourse. Number of fluency disruptions and production rate are inextricably linked. A slower rate generally indicates that more non-communicative pauses and fillers occupy output time. In keeping with the findings of Field and colleagues (2000), both groups increase their production rate across retellings, but unlike the findings of Field et al., 2000 and Howell et al., 1994, the TLE group remains consistently slower in our data. While the disparity between groups was not evident after three trials (Field et al., 2000), plausibly the increased number of elicitations in the present study has led to a widening and clarification of this effect.

While dysfluencies, volume, and rate of output are saliently perceptible features from a listener’ s perspective, it is cohesion and coherence which affect the quality of discourse, lead to the listener’ s impression of circumstantiality, and reflect an inefficient linguistic system in TLE. Discourse production relies heavily on working memory (see Stretton & Thompson, 2012 for review), and disturbances impede the capacity to maintain the current discourse representation in a manner that allows for active revision, updating, and ultimately refinement (Bell et al., 2003; Canoz & Vion, 1994; Daneman, 1991; Field et al., 2000; Fletcher & Henson, 2001; Hartley & Jensen, 1991; Howell et al., 1994; Johns et al., 2008; Levelt, 1989). Since cohesion relies largely on an individual’ s capacity to monitor their own output and the listener’ s understanding simultaneously, capacity limitations and failures of working memory ultimately manifest as cohesion disturbances, particularly in the form of ambiguous, incomplete, or missing personal referents. Referents maintain clear and unambiguous links to previously introduced content (Halliday & Hasan, 1976); for example, *Eliza went swimming. <u>She</u> was exhausted*. Losing track of this mental representation leads to cohesion errors, signifying a disruption in the speaker’ s understanding of connections between linguistic elements (Bloom, 1994). Repetitive statements in TLE also reflect failures of attention and working memory which lead to disturbances in forming, retaining, and monitoring dynamic discourse representation. Repetitions might serve as attempts to compensate for disrupted cohesion by clarifying referents and narrative more broadly after losing track of what has been said. Individuals with TLE and controls equally communicated core relevant content, but coherence in TLE was impeded by the inclusion of fewer novel units, and consistently more non-progression errors, including extraneous or repetitive content. Similar coherence disturbances are reported in patients with damage to the right hemisphere (Abusamra et al., 2009; Joanette et al., 1986; Johns et al., 2008; Marini, 2012).

While these findings suggest that cohesion and coherence disturbances in TLE are underpinned by a capacity limitation, the role of pragmatic dysfunction is worth considering. Rao and colleagues (1992) postulated that circumstantiality in TLE reflects a need to maintain social contact. Peripheral remarks, including topic shifts and personalised statements such as “I had a toy horse as a kid” might represent dysfunctional social cognition (Broicher et al., 2012; Garcia & Joanette, 1994; Schacher et al., 2006) and an attempt to compensate for reduced novelty and informativeness (Field et al., 2000). Extraneous and tangential details are more common in TLE than controls and have been previously reported (Lomlomdjian et al., 2017). To take an example from our participant with TLE at Trial 4, “Well, I’ ll call the people their names. This is Roy on- on his nice horse…and then a little boy- oh his name is um David. He comes with his toy horse”. This excerpt indicates an elaboration with extraneous details that do not directly progress the narrative. While all participants in re-elicitation research might misinterpret the request as having done ‘something wrong’, individuals with TLE are more likely to compensate by producing *more* output in a bid to clarify content, create novelty, and maintain the examiner’ s interest, manifesting in their “sticky” interpersonal style (Rao et al., 1992), in other words, a heightened drive to ‘hold the floor’. Healthy controls are not only able to compress discourse but might also have a greater understanding of the pragmatics of this interaction. The need for social desirability, in the service of pragmatic goals, could also in part explain repetitiveness in TLE if they believe they have been misunderstood, unclear, or uninformative (Bloom, 1994). Pedantic attention to detail can be conceptualised as a pragmatic issue, where there is a tendency to make erroneous judgements about what is and is not relevant and this effect deepens with further elicitations. Discourse production and the mental representation of discourse reflect more than the text and its lexical choices; they encapsulate the pragmatic demand to consider contextual factors and extra-linguistic domains such as Theory of Mind (ToM), as well as information about the conversational partner (Hagoort & Van Berkum, 2007; Xu et al., 2005). The resultant discourse is therefore a product of all components of the interaction, including its sociolinguistic and situational context, such as repeated elicitation (Johnson-Laird, 1983; van Dijk & Kintsch, 1983; Xu et al., 2005).

In this population, word-findings difficulties and microlinguistic impairments such as fluency disruptions, duration of pauses, and production rate are typically attributed to interictal disturbances to language function and our understanding of the direct impact of seizure burden and ASM usage on processing speed via network disturbances (Englot et al., 2010; Kwan &Brodie, 2001). While seizure burden positively correlated with core discourse metrics at the final trial, there was no significant effect on linear mixed models. The implication of this finding is that macrolinguistic disturbances in language that are seen in TLE occur over and above the immediate consequences of seizure and interictal activity, and instead relate to the syndrome of TLE more broadly. Various aspects of the neural network subserving ToM might be impacted in TLE, including medial prefrontal cortices, superior temporal sulcus, bilateral temporal poles, and amygdala (Broicher et al., 2012; Giovagnoli et al., 2011; Giovagnoli, Parente, Villani, Franceschetti, & Spreafico, 2013; Schacher et al., 2006).

This study is concerned with high-level linguistic function which is not strictly represented in a simplistic modular or focal fashion. While more fundamental aspects of cognitive-linguistic function can be localised or lateralised, this is not necessarily the expectation for higher-order language, as a higher cortical function. Functions as complex as this are seldom as localised as lateralised as one might think, and instead relate to widely distributed networks. In light of this, right and left TLE participants were considered as a single diagnostic group. While this might sound counterintuitive, there was no sense in which the groups were distinguishable. This approach was supported by our analyses which indicated that there were no differences in discourse outcomes when considered as separate groups or collectively, and ultimately that treating them as a single group was a robust and practical choice. As a preliminary enquiry to address naturalistic output with detailed discourse analysis, this study generated a very large corpus of data for each participant rather than dealing with single, disparate data points. This allowed the examination of complex functions. Future research with more evenly distributed groups might be useful to confirm the veracity of these findings or to determine possible subtle differences between these groups.

This study extends seminal work on narrative discourse in TLE. There is a clear hierarchy in the emergence of a pragmatic disorder from network disruption, beginning with effects on transformational grammar, and extending to the expectations of the interlocutor and social interaction more generally. While on initial observation the number of elicitations could be perceived as excessive and potentially confounding, it in fact represents exactly what we seek to understand, that is, language in practice.

## Data Availability

All data produced in the present study are available upon reasonable request to the authors

## Acknowledgement of funding

This work was financially supported by the Australian Government Research Training Program Scholarship (Stipend and Fee offset) awarded by the Australian Commonwealth Government and the University of Melbourne to the first author.

## Author Contributions

**Fiore D’ Aprano:** Conceptualisation (equal); Methodology (equal); Investigation (lead); Data Curation (lead); Formal Analysis (lead); Project Administration (lead); Visualisation (lead); Writing – Original Draft Preparation (lead); Writing – Review & Editing (equal).

**Charles B. Malpas:** Conceptualisation (equal); Methodology (equal); Formal Analysis (supporting); Resources (equal); Writing – Review & Editing (equal), Supervision (equal)

**Stefanie E. Roberts:** Data curation (supporting); Methodology (equal); Validation (lead)

**Michael M. Saling:** Conceptualisation (equal); Methodology (equal); Resources (equal); Writing – Review & Editing (equal), Supervision (equal)

