## Appendix A for "Verbosity with retelling: narrative discourse production in temporal lobe epilepsy"

**Verb Generation Task (VGT) development**

The Verb Generation Task (VGT) was specifically developed for the purpose of this study to consider verb-retrieval deficits in this population. This task was used to examine the retrieval of verbs on the basis of visual stimuli. For this task, participants are shown a series of 60 black and white line drawings taken from Snodgrass and Vanderwart stimuli (Snodgrass & Vanderwart, 1980). These were normed for name agreement, image agreement, familiarity, and visual complexity (Snodgrass & Vanderwart, 1980). On the basis of a psycholinguistic database (Wilson, 1988), images were then selected on their ratings of concreteness to include an equal number of items that had concreteness ratings above and below 600, corresponding with items that were considered concrete and abstract, respectively. Items were also selected to ensure that there was an equal distribution of items with high and low verb agreement. The images were then collated in a randomised order, and the order of presentation was identical for all participants. Participants were asked to provide the associated verb that first comes to mind in response to each visual stimulus. Alternative morphological forms were accepted, for example a target infinitive form "sleep" would also allow "to sleep" "sleeping" and "sleeps". Responses were recorded based on whether the correct dominant verb was provided, or whether a non-dominant correct response was provided. These included all responses that were verbs that could plausibly relate to the stimulus. Incorrect responses referred to those that either were not verbs, or did not relate to the stimulus in a reasonable way. The variables of interest relate to the number of correct dominant responses, the number of correct non-dominant response. Their response latencies were also recorded to examine the total latency across all items, and a mean latency metric.
