## Appendix C for "Verbosity with retelling: narrative discourse production in temporal lobe epilepsy"

**Table 1**

*Sample characteristics for neuropsychological variables*

| | Control (*n* =14) | | TLE (*n* =15) | | *p* | *r* |
|---|---|---|---|---|---|---|
| Variable | Median *(Q1,Q3)* | Range | Median *(Q1,Q3)* | Range | | |
| TOPF[a] | 0.75 (-0.02, 1.33) | 2.25 | 0.25 (-0.67, 0.67) | 3.00 | 0.06 | 0.41 |
| WASI-II $_{\text{Matrix Reasoning}}$ | 0.40 (0.00, 0.92) | 6.50 | 0.00 (-0.33, 0.55) | 4.50 | 0.40 | 0.19 |
| WASI-II $_{\text{Vocabulary}}$ | 1.00 (-0.18, 1.25) | 3.50 | 0.55 (0.14, 0.67) | 3.67 | 0.20 | 0.29 |
| WASI-II $_{\text{FSIQ}}$ | 0.61 (-0.25, 0.75) | 3.17 | 0.25 (-0.42, 0.55) | 4.50 | 0.26 | 0.25 |
| WAIS-IV Digits Forward[a] | 0.34 (-0.67, 0.92) | 3.67 | -0.33 (-0.33, 0.17) | 2.33 | 0.47 | 0.16 |
| WAIS-IV Digits Backward[a] | 0.00 (-0.67, 0.33) | 3.34 | 0.00 (-0.33, 0.50) | 2.66 | 0.98 | 0.01 |
| WMS-IV Logical Memory I[a] | 0.67 (-0.16, 1.33) | 4.00 | 0.33 (-0.17, 1.00) | 2.33 | 0.58 | 0.12 |
| WMS-IV Logical Memory II[a] | 1.00 (0.00, 1.00) | 3.66 | 0.33 (-0.33, 1.00) | 2.67 | 0.48 | 0.16 |
| RAVLT $_{\text{Total}}$[a] | 0.59 (-0.09, 1.52) | 3.57 | 0.18 (-0.15, 1.15) | 3.01 | 0.62 | 0.11 |
| RAVLT $_{\text{Post-Interference Recall}}$[a] | 0.22 (-0.58, 1.10) | 3.01 | 0.22 (-0.86, 0.89) | 3.76 | 0.65 | 0.10 |
| RAVLT $_{\text{Delayed Recall}}$[a] | 0.28 (-0.46, 1.26) | 4.34 | 0.32 (-0.77, 0.69) | 3.91 | 0.65 | 0.10 |
| WMS-R Easy^ | 12.00 (11.00, 12.00) | 3.00 | 11.00 (10.00, 11.50) | 3.00 | 0.18 | 0.28 |
| WMS-R Hard^[a] | 8.50 (6.25, 10.00) | 10.00 | 7.00 (6.00, 8.50) | 9.00 | 0.21 | 0.28 |
| WMS-R Easy Delay^ | 4.00 (4.00, 4.00) | 0.00 | 4.00 (4.00, 4.00) | 1.00 | 0.37 | 0.07 |
| WMS-R Hard Delay^ | 4.00 (3.25, 4.00) | 4.00 | 4.00 (3.00, 4.00) | 2.00 | 0.60 | 0.10 |
| Victoria Stroop $_{\text{Dots}}$[a] | -0.83 (-1.25, -0.33) | 2.33 | -1.33 (-1.33, -1.00) | 3.34 | 0.12 | 0.34 |
| Victoria Stroop $_{\text{Words}}$[a] | -0.50 (-0.92, -0.33) | 1.67 | -0.67 (-1.33, -0.33) | 2.33 | 0.31 | 0.22 |
| Victoria Stroop $_{\text{Colour}}$[a] | 0.00 (-0.92, 0.67) | 4.00 | 0.00 (-0.67, 0.33) | 2.00 | 0.60 | 0.12 |
| Victoria Stoop $_{\text{Interference}}$[a] | 0.83 (-0.83, 1.00) | 3.33 | 1.00 (0.50, 1.33) | 2.00 | 0.41 | 0.18 |
| BNT $_{\text{Total}}$[a] | -0.05 (-0.67, 0.55) | 3.03 | -0.74 (-1.65, 0.13) | 3.84 | 0.12 | 0.35 |
| BNT $_{\text{TOT States}}$^[a] | 17.00 (13.75, 23.50) | 22.00 | 24.00 (21.00, 27.00) | 17.00 | 0.02* | 0.48 |
| BNT $_{\text{Proportion TOT}}$^[a] | 0.28 (0.23, 0.40) | 0.37 | 0.40 (0.35, 0.44) | 0.20 | 0.08 | 0.39 |
| OLR (COWAT)[a] | -0.53 (-0.98, -0.04) | 4.01 | -1.17 (-1.58, 0.16) | 4.29 | 0.29 | 0.23 |
| Animals[a] | 0.67 (-0.02, 1.63) | 3.34 | -0.35 (-0.61, 0.30) | 2.39 | 0.0099** | 0.57 |
| ANT $_{\text{Total}}$^ | 48.00 (48.00, 49.00) | 5.00 | 47.00 (45.00, 49.00) | 9.00 | 0.29 | 0.23 |
| ANT $_{\text{Latency}}$^[a] | 64.00 (59.50, 69.00) | 23.00 | 84.00 (68.50, 92.00) | 60.00 | 0.02* | 0.53 |
| ANT $_{\text{TOT States}}$^[a] | 11.50 (9.00, 14.50) | 18.00 | 21.00 (13.00, 27.00) | 29.00 | 0.03* | 0.47 |
| ANT $_{\text{Proportion TOT}}$^[a] | 0.23 (0.18, 0.29) | 0.36 | 0.42 (0.26, 0.54) | 0.58 | 0.03* | 0.47 |
| WFD Frequency Rating^[a] | 2.50 (1.25, 3.00) | 3.00 | 3.00 (3.00, 4.00) | 5.00 | 0.04* | 0.45 |
| WFD Distress Rating^ | 1.00 (1.00, 1.75) | 3.00 | 4.00 (3.00, 6.00) | 6.00 | 0.0006** | 0.74 |
| VGT $_{\text{Correct Dominant}}$^ | 36.50 (34.25, 42.75) | 20.00 | 36.00 (32.00, 38.00) | 22.00 | 0.34 | 0.21 |
| VGT $_{\text{Correct Other}}$^ | 18.00 (15.00, 21.00) | 15.00 | 19.00 (17.00, 21.25) | 14.00 | 0.46 | 0.17 |
| VGT $_{\text{Latency}}$^ | 184.50 (164.25, 196.75) | 125.00 | 213.00 (183.50, 308.00) | 323.00 | 0.03* | 0.48 |
| HADS $_{\text{Anxiety}}$^[a] | 7.00 (5.25, 8.50) | 10.00 | 7.00 (6.50, 8.50) | 10.00 | 0.58 | 0.12 |
| HADS $_{\text{Depression}}$^ | 2.00 (1.00, 3.00) | 9.00 | 4.00 (4.00, 5.50) | 16.00 | 0.022* | 0.50 |

*Note.* TLE = Temporal Lobe Epilepsy; TOPF = Test of Premorbid Functioning; WASI-II = Wechsler Abbreviated Scale of Intelligence, Second Edition; FSIQ = Full Scale Intelligence Quotient; WAIS-IV = Wechsler Adult Intelligence Scale, Fourth Edition; WMS-IV = Wechsler Memory Scale, Fourth Edition; RAVLT = Rey Auditory Verbal Learning Test; WMS-R = Wechsler Memory Scale–Revised; BNT = Boston Naming Test; TOT = Tip-of-the-tongue; OLR (COWAT) = Orthographic Lexical Retrieval (Controlled Oral Word Association Test); WFD = Word Finding Difficulty; VGT = Verb Generation Task; ANT = Auditory Naming Task; HADS = Hospital Anxiety and Depression Scale. Represented as z-scores where normative data was available, raw data are indicated by^. Latencies are expressed in seconds. Group differences computed using Mann-Whitney U test, effect sizes *r*, where * = p <.05, ** = p <.01. [a]Suggests that data does not violate assumptions of normality on Shapiro-Wilk.

**Table 2**

*Group differences on discourse variables across trials*

| Discourse Measure | Trial 1 | | Trial 2 | | Trial 3 | | Trial 4 | | Trial 5 | |
|---|---|---|---|---|---|---|---|---|---|---|
| | *MD* [95% CI] | *p* [RBC] | *MD* [95% CI] | *p* [RBC] | *MD* [95% CI] | *p* [RBC] | *MD* [95% CI] | *p* [RBC] | *MD* [95% CI] | *p* [RBC] |
| Sample length | 3.91 [-52.00, 60.00] | 0.97 [0.01] | -4.00 [-52.00, 38.00] | 0.86 [0.04] | -18.00 [-68.00, 24.00] | 0.36 [0.20] | -29.80 [-102.00, 6.00] | 0.10 [0.36] | -30.35 [-96.00, 14.00] | 0.19 [0.29] |
| Spontaneous duration | -13.76 [-46.02, 16.17] | 0.26 [0.25] | -13.23 [-35.92, 6.85] | 0.16 [0.31] | -15.99 [-40.87, 2.24] | 0.10 [0.36] | -21.58 [-44.29, 0.95] | 0.057 [0.42] | -24.47 [-44.49, -2.98] | 0.018 [0.51]*† |
| Pause duration | -10.40 [-28.28, -0.63] | 0.033 [0.47]* | -6.89 [-15.99, -1.55] | 0.01 [0.53]* | -9.53 [-21.93, -3.55] | 0.001 [0.68]*† | -7.90 [-23.33, -2.51] | 0.0060 [0.59]* | -10.99 [-18.46, -6.65] | 0.0001 [0.78]*† |
| Duration (excluding pauses) | -2.83 [-19.80, 15.36] | 0.88 [0.04] | -4.71 [-19.26, 9.42] | 0.51 [0.15] | -5.96 [-22.02, 5.88] | 0.39 [0.19] | -8.42 [-25.84, 5.19] | 0.19 [0.30] | -11.30 [-28.32, 2.64] | 0.10 [0.36] |
| Production rate (words/second) | 0.42 [0.12, 0.72] | 0.011 [0.56]*† | 0.28 [0.00, 0.61] | 0.049 [0.43]* | 0.43 [0.08, 0.68] | 0.013 [0.55]*† | 0.26 [-0.04, 0.52] | 0.097 [0.37] | 0.42 [0.12, 0.67] | 0.0049 [0.62]*† |
| Total statements | -1.00 [-9.00, 7.00] | 0.78 [0.07] | -2.00 [-7.00, 5.00] | 0.60 [0.12] | -4.00 [-12.00, 2.00] | 0.22 [0.27] | -4.00 [-15.00, 1.00] | 0.16 [0.31] | -6.43 [-14.00, -0.00] | 0.044 [0.44]* |
| Fluency disruptors^ | -0.09 [-0.15, -0.03] | 0.006 [0.60]*† | -0.06 [-0.13, -0.00] | 0.042 [0.45]* | -0.08 [-0.14, -0.02] | 0.009 [0.57]*† | -0.07 [-0.12, -0.02] | 0.015 [0.53]* | -0.06 [-0.11, -0.01] | 0.018 [0.52]*† |
| Clarity disruptors^ | 0.01 [-0.01, 0.02] | 0.40 [0.19] | 0.00 [-0.01, 0.01] | 0.76 [0.07] | 0.00 [-0.01, 0.01] | 0.48 [0.15] | 0.00 [-0.01, 0.01] | 0.99 [0.00] | -0.00 [-0.01, 0.01] | 0.64 [0.10] |
| False starts^ | -0.01 [-0.03, 0.02] | 0.44 [0.17] | -0.00 [-0.05, 0.01] | 0.61 [0.11] | -0.00 [-0.03, 0.02] | 0.83 [0.05] | -0.01 [-0.04, 0.01] | 0.33 [0.21] | 0.01 [-0.00, 0.03] | 0.14 [0.32] |
| Fillers (non-grammatical)^ | -0.00 [-0.01, 0.01] | 0.51 [0.14] | -0.00 [-0.01, 0.01] | 0.96 [0.01] | -0.01 [-0.02, 0.01] | 0.40 [0.19] | 0.00 [-0.01, 0.02] | 0.77 [0.07] | 0.01 [-0.00, 0.01] | 0.20 [0.27] |
| Pauses (non-grammatical)^ | -0.05 [-0.08, -0.03] | 0.0006 [0.75]*† | -0.03 [-0.06, -0.01] | 0.018 [0.52]* | -0.05 [-0.07, -0.02] | 0.0008 [0.73]*† | -0.03 [-0.06, -0.00] | 0.035 [0.46]* | -0.05 [-0.07, -0.02] | 0.004 [0.63]*† |
| Hesitations (non-grammatical)^ | -0.06 [-0.11, -0.03] | 0.0023 [0.67]*† | -0.04 [-0.07, -0.01] | 0.032 [0.47]* | -0.06 [-0.09, -0.02] | 0.0009 [0.72]*† | -0.04 [-0.06, -0.00] | 0.049 [0.42]* | -0.04 [-0.08, -0.01] | 0.013 [0.54]*† |
| Non-grammatical hesitations to all^ | -0.10 [-0.19, -0.02] | 0.024 [0.50]* | -0.02 [-0.14, 0.08] | 0.66 [0.10] | -0.08 [-0.21, 0.03] | 0.13 [0.33] | -0.05 [-0.13, 0.07] | 0.35 [0.21] | -0.06 [-0.18, 0.05] | 0.24 [0.26] |
| Other referents^ | -0.01 [-0.02, -0.00] | 0.027 [0.47]* | -0.02 [-0.03, -0.00] | 0.0082 [0.57]* | -0.02 [-0.04, -0.01] | 0.0049 [0.61]*† | -0.00 [-0.02, 0.01] | 0.38 [0.19] | -0.01 [-0.02, -0.00] | 0.021 [0.49]*† |
| Disrupted Cohesion | -7.00 [-11.00, -1.00] | 0.0059 [0.60]*† | -3.00 [-8.00, -1.00] | 0.0057 [0.60]* | -3.00 [-6.00, -1.00] | 0.0085 [0.58]*† | -3.00 [-6.00, -1.00] | 0.021 [0.50]* | -4.00 [-8.00, -1.00] | 0.0028 [0.65]*† |
| Core Propositions | 1.20 [-2.00, 4.00] | 0.50 [0.15] | 1.00 [-1.00, 4.00] | 0.29 [0.23] | 1.00 [-1.00, 3.00] | 0.34 [0.21] | -0.00 [-3.00, 3.00] | 0.99 [0.00] | 1.00 [-2.00, 3.00] | 0.74 [0.08] |
| Task-on novel units# | 0.14 [0.03, 0.25] | 0.025 [0.50]* | 0.07 [-0.04, 0.20] | 0.11 [0.35] | 0.10 [0.01, 0.19] | 0.040 [0.45]* | 0.13 [0.03, 0.24] | 0.0063 [0.60]* | 0.17 [0.08, 0.25] | 0.0011 [0.72]*† |
| Non-progression units# | -0.11 [-0.23, 0.01] | 0.060 [0.41] | -0.07 [-0.20, 0.07] | 0.24 [0.26] | -0.10 [-0.20, 0.00] | 0.043 [0.42]* | -0.14 [-0.24, -0.03] | 0.0063 [0.60]*† | -0.17 [-0.25, -0.09] | 0.0001 [0.83]*† |
| Non-progression to task-on novel units | -0.26 [-0.60, -0.01] | 0.040 [0.45]* | -0.10 [-0.39, 0.09] | 0.23 [0.27] | -0.17 [-0.36, 0.01] | 0.049 [0.41]* | -0.24 [-0.45, -0.05] | 0.0063 [0.60]*† | -0.30 [-0.51, -0.15] | 0.0002 [0.82]*† |
| Syntactic Simplicity^ | -0.01 [-0.04, 0.01] | 0.21 [0.28] | -0.01 [-0.05, 0.02] | 0.47 [0.16] | -0.01 [-0.04, 0.00] | 0.059 [0.41] | 0.01 [-0.01, 0.03] | 0.29 [0.23] | -0.01 [-0.04, 0.01] | 0.37 [0.20] |

*Note.* TLE = Temporal Lobe Epilepsy; MD = Mean Difference where Control – TLE, CI = Confidence Interval. Metrics marked ^ are expressed relative to sample length, # are expressed relative to total statements. Group differences computed using Mann-Whitney U test, effect sizes expressed as Rank Biserial Correlation, where * = *p* <.05, † = significance holds on false detection rate (FDR) correction.
